# Assessment of Attitude and Hesitancy Towards Covid-19 Vaccine among Hepatitis B and C Patients in Pakistan

**DOI:** 10.1101/2022.11.23.22282686

**Authors:** Farheen Shafiq, Mahreen ul Hassan, Sadia Butt, Sadia Sidique, Nazia Akbar, Azra, Irshad Rehman

**Author notes:** **Correspondence: Dr. Mahreen ul Hassan**, Contact no.03348483041, 78 Western Bank, Firth Court, Department of Bioscience, University of Sheffield, UK, S10 2TN, Sheffield, South Yorkshire, United Kingdom.

## Abstract

**OBJECTIVE:** The research aimed to evaluate the attitude and perceptions towards the covid-19 vaccine among Hepatitis B and C patients in Peshawar, Khyber Pakhtunkhwa, Muzaffarabad, Azad Kashmir, Pakistan.

**METHODS:** A survey-based study was adopted to evaluate the attitude of Hepatitis B and C patients towards immunization against covid-19 in Peshawar (KPK) and Muzaffarabad (AJK) cities of Pakistan. The study continued from January 2020 to February 2021. Participants were also assessed for their perception towards covid-19 vaccination.

**RESULTS:** A total of 839 (33.6%) individuals participated in the study. About 52 % of Hepatitis B patients were immunized against Covid-19, whereas the number of Hepatitis C patients was recorded at around 48%. About 53.7 % of participants refused to get the vaccine without any reason. About 63.2% of patients showed concern about the insufficient data available on the vaccine safety and efficacy published by the Public Health Department. Individuals with higher education were observed to be more open towards vaccination then those without a formal education. More than half of the participants (61.5 %) were concerned about the interference of the vaccine with their hepatitis treatment whereas 54.7 % patients refused vaccine because of a poor liver condition.

**CONCLUSIONS:** The data indicated that limited data availability regarding the vaccine efficacy in viral hepatitis patients and negative attitudes of people toward covid-19 vaccination is the main cause of Covid-19 vaccination refusal among hepatitis B and C patients.

**DESCRIPTORS:** Hepatitis B, Hepatitis C, covid-19, immunization, vaccine refusal, Pakistan.

## INTRODUCTION

Severe Acute Respiratory Syndrome Coronavirus 2 (SARS-CoV-2), the 7th human coronavirus, was reported in Wuhan, Hubei province, China, in January 2020^1-2^. In early 2020, the World Health Organization (WHO) declared it a pandemic and requested the world to take strict measures to control the disease as there was no effective treatment available, and the death toll was rising every hour across the world^3^.

The SARS-CoV-2 is an airborne virus which can be transmitted through air therefore effective preventive measures like social distancing, frequent handwash and wearing a mask in the absence of a valid treatment or vaccine to control the disease was imperative^4^.

Patients with covid-19 may have abnormal liver function or hepatic dysregulation, which may identify as cholestasis, hepatitis, or both. In an early study from China, increased total bilirubin was observed in 18 (18%) patients and higher serum alanine aminotransferase (ALT) was recorded in 28 (28%) of 99 individuals with covid-19^5^.

It is suggested that SARS-CoV-2 may exacerbate liver damage in individuals with chronic viral hepatitis^6^. According to Xu et al.^7^ (2020), liver dysfunction occurs more often and more severely as covid-19 progresses. Liver function tests are typically normal or somewhat raised in the early stages of SARS-CoV-2 infection. Histological examination of the liver reveals syncytial multinuclear hepatocytes, microvascular steatosis, and little lobular and portal activity.

The cytokine storm caused by the progression of severe covid-19 cases requiring strong immunosuppressive therapy has raised the problem of recurrence of HBV infections in the patients with a history of HBV^8^.

In a study conducted by Ronderos et al.^9^ (2021), patients with covid-19 having history of hepatitis C virus (HCV) or seropositivity for HCV infection have an increased susceptibility to SARS-CoV-2 infection, which is a strong predictor of in-hospital mortality regardless of preexisting conditions, laboratory results upon admission, or the severity of covid-19-induced liver injury.

With an estimated 9.8 million chronic HCV patients, Pakistan has the second-highest burden of HCV infection in the world^10^. Unscreened blood transfusions, a history of hospitalization, dental work, tattooing, body piercing, the use of injections, and a history of surgery are the main risk factors for the transmission of HCV^11^. According to government statistics, around 2.5% of Pakistan’s population was infected with HBV in 2008. There was a 5% rise in HCV-related mortality and an 8% increase in HBV-related deaths between 2015 and 2019^10^.

According to World Health Organization vaccination is an easy, safe, and efficient way to avoid infection especially in the absence of an effective therapy as in the case of covid-19^12^. Vaccination is the most significant medical intervention in the human history which has largely eradicated the infectious diseases that formerly claimed the lives of millions of people^13^. Currently, more than 30 infectious diseases could be avoided with the use of vaccinations^14^. According to an estimation, the immunization against covid-19 have prevented thousands of deaths and saved millions of people from getting infection in England and United States till June 2021^15-16^.

Pakistan is also badly affected by covid-19 pandemic. Currently there are 1,573,922 confirmed covid-19 cases from which 2,928 are marked as active in different areas of Pakistan and about 30,625 causalities have been observed from 2020 to date^17^.

The Chinese government sent more than 500,000 doses of the Chinese Sinopharm vaccine to Pakistan in February 2021, and the Pakistani government opted to prioritize immunizing frontline healthcare workers^18^. Pakistan initially restricted immunization to those aged 18 and older before expanding the program. The government was placing a higher priority on heavily populated metropolitan centers and “hotspot” locations, where the coronavirus transmits more rapidly than in more distant rural communities^19^. It is estimated that about 69.9 % population in Pakistan has taken at least on dose of covid-19 vaccine. The number is not very discouraging but nature of disease and its transmission along with the emergence of new Corona virus variants every 6 month makes it crucial to get the entire population immunized in limited time to control the disease^17^.

Vaccine hesitancy is one of the top 10 global health problems, according to the World Health Organization^20^. Fear of the covid-19 vaccination is a growing problem that Pakistan is also dealing with. It is one of the nations with lowest vaccination rates^21^. Myths such as the vaccine causes sterility, the pandemic is a foreign conspiracy, or the virus is harmless are common contributors to the hesitation towards covid-19 vaccination^22^.

In current study have sought to understand the current level of awareness of covid-19 transmission, symptoms, and preventive measures specifically among Hepatitis B and C patients. The study also aimed to identify the risk variables that contribute to ambivalence towards covid-19 vaccination among hepatitis patients residing in KPK and AJK, Pakistan.

## METHODOLOGY

### Study Location and Settings

Survey-based research was conducted in Peshawar and Muzaffarabad from January 2020 to February 2021. A questionnaire was prepared for the study. An epidemiologist, field surveyor, and health care practitioner assisted in the development of a questionnaire. To calculate the reliability score and Cronbach alpha, the questionnaire was pretested in a pilot study.

### Sample Size and Exclusion Criteria

For the evaluation of the knowledge and perception of the hepatitis B and C patients toward covid-19 and the vaccination program the questionnaire was distributed throughout the community where these patients were easily accessible. Hepatitis B and C patients age 18 years and above were deemed eligible. Participants who refused to provide any information or who were temporary residents were barred from participating. The questionnaire was created to identify potential challenges to covid-19 immunization.

### Designing and Validating the Questionnaire

Following a thorough review of the relevant literature, a questionnaire was designed. It was composed in English, then translated into Urdu and Pushto for participants before translated back to English for analysis. The questionnaire was divided into two sections, each with 16 questions. Section one had ten questions that assessed demographic data such as age, gender, race, income, marital status, and so on. The next section included 16 questions designed to test participants’ knowledge and perceptions about covid-19 and its vaccination.

### Statistical Analysis

Data evaluation was performed through SPSSv21 statistical package.

### Ethical Approval and Consent of Participants

This study was approved by the Research and Ethics Committee of Shaheed Benazir Bhutto Women University Peshawar, Pakistan. A filled and signed informed consent form was collected from each participant before the participation. All the information of the participants is kept confidential.

## RESULTS

### Demographic Details of the Respondents

A total of 839 responses were collected, of which 36.1% (303) were female and 63.9% (536) were from male respondents. Table 1 displays the demographic data (age, gender, place of residence, educational background, occupation, and monthly income) of the survey respondents. About 48% responses were collected from Hepatitis B patients whereas the response rate of Hepatitis C patients was 52 %. Age-wise most of the patients were young (57.1%) while it was observed that the number of respondents were mostly male (63.9 %) that could be due to the cultural barriers in the area. The data generated through this study revealed the about 35.5% hepatitis patients were still unvaccinated due to various reasons.

**Table 1.**
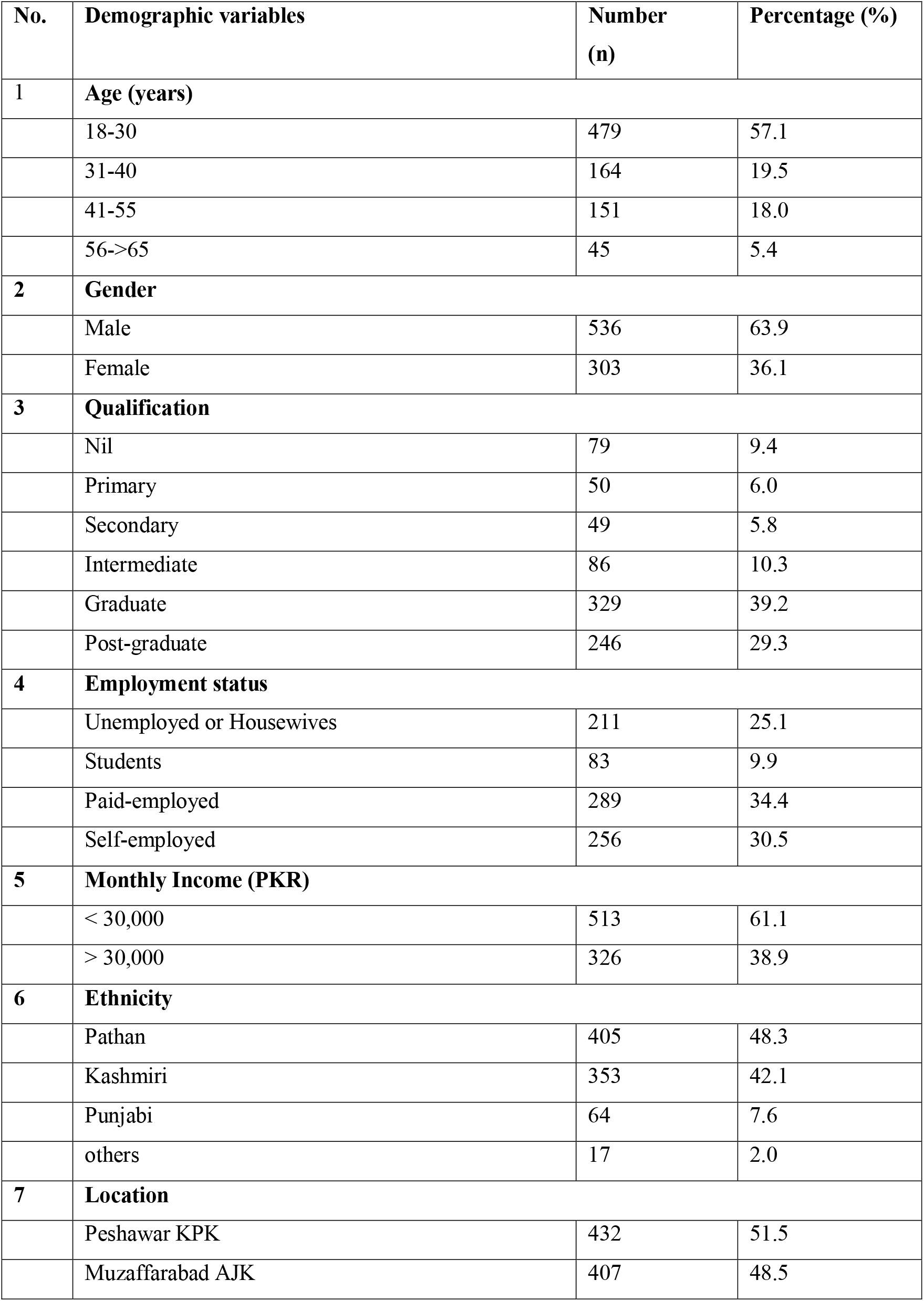
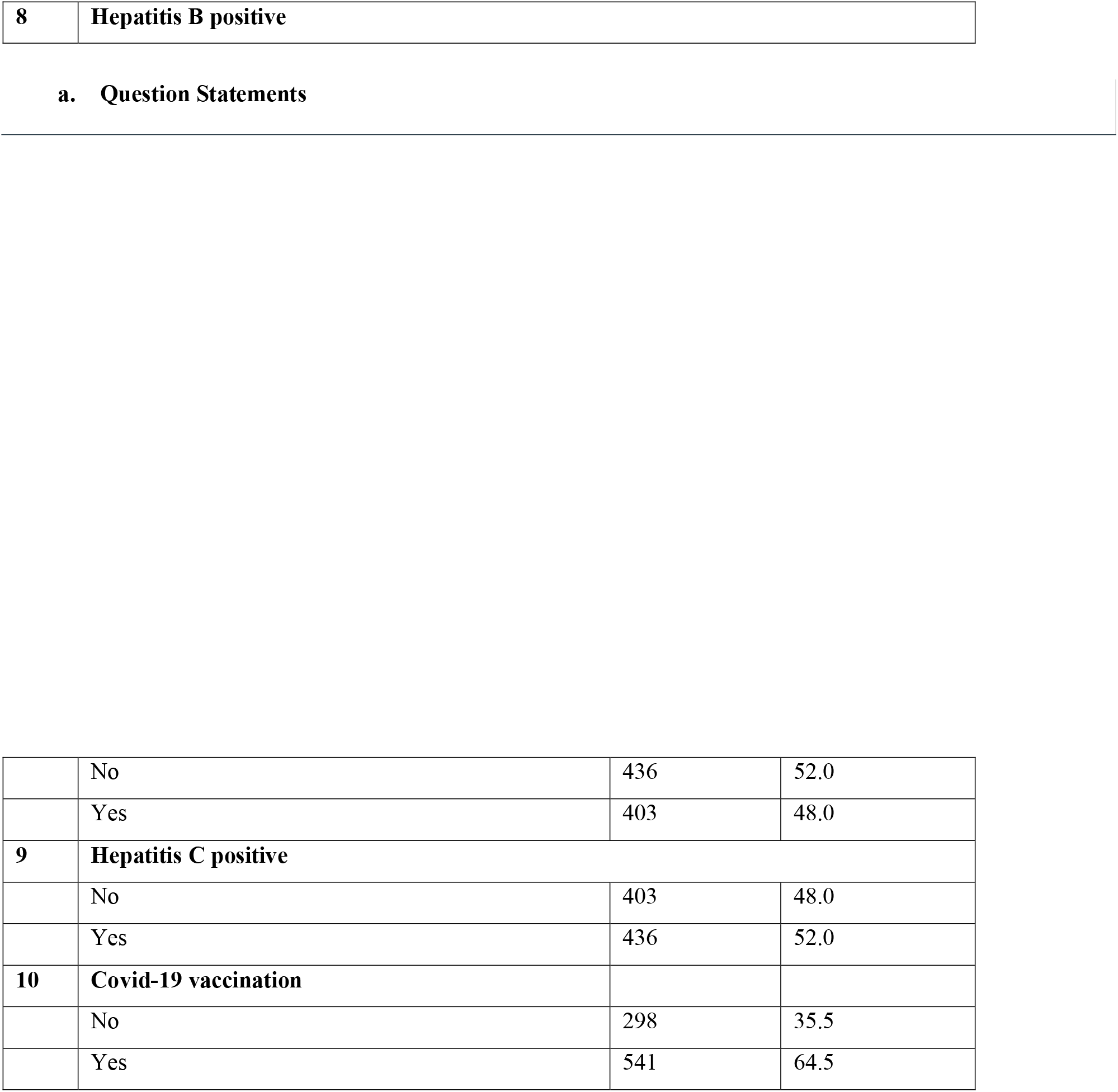
shows demographic data frequencies of the participants. (n = 839).

### Approved Vaccines in Pakistan

In Pakistan, every person above age 18 was bound to get one of the approved Covid-19 vaccines to minimize the devastating effects of pandemic according to National Action Plan for Corona virus disease (covid-19) Pakistan^a^ (Table S1). For the purpose, Government of Pakistan worked tirelessly making the best vaccines widely available to the public. As a result, about 10% of the population received the first by the mid of year 2021. The information presented in Table 6 was generated as a secondary data by utilizing all the reliable resources available e.g., National Institute of Health Pakistan^b^, WHO^c^ and dedicated website for covid-19 related information by the Ministry of National Health Services Pakistan^d^. Likewise, it was advised that everyone with a known underlying disease, including those with hepatitis, immunize themselves against covid-19.

It was observed during the study that around 65 % participants were fully immunized (Figure 1a), and rest of the participants refused after taking first dose to immunize due to some reasons (Figure 1b).

**Figure 2.**
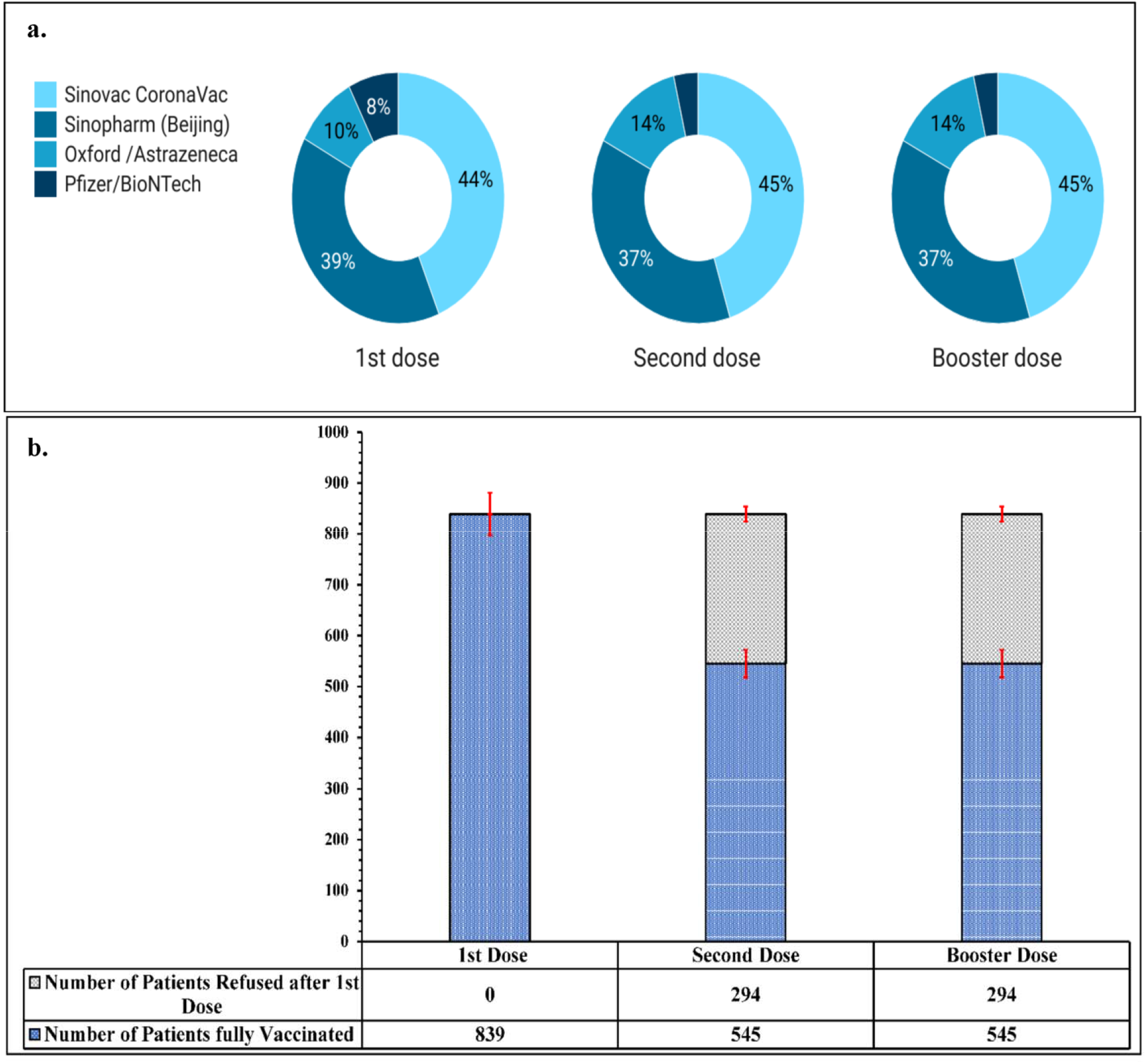
a. No. of patients receiving Covid vaccines from approved pharmaceuticals. b. No. of Patients refused to get immunized.

The main objective of this study was to evaluate the hepatitis patients’ awareness of the covid-19 vaccination and the fears that were keeping them from receiving the vaccine. For the purpose a derailed interview was arranged with each participant and their answer were recorded for further analysis.

### Attitude and Perceptions towards covid-19 vaccination

The questionnaire was split in two parts i.e., part A (Table 3a) where patients were assessed why they chose to get immunized against covid-19. Part B (Table 3b) was specifically designed for the patients who refused to take covid vaccine.

**Table 2.** -Commercially available Vaccines approved by the Government of Pakistan.

| Approved Vaccines | Type of vaccine |
| --- | --- |
| Moderna mRNA 1273 | RNA |
| Pfizer/BioNTech BNT 162b2 | RNA |
| Consino Ad5-NCov | Non-Replicating viral Vector |
| Gamaleya Spuntik V | Non-Replicating viral Vector |
| Oxford /Astrazeneca AZD1222 | Non-Replicating viral Vector |

**Table 3a.**
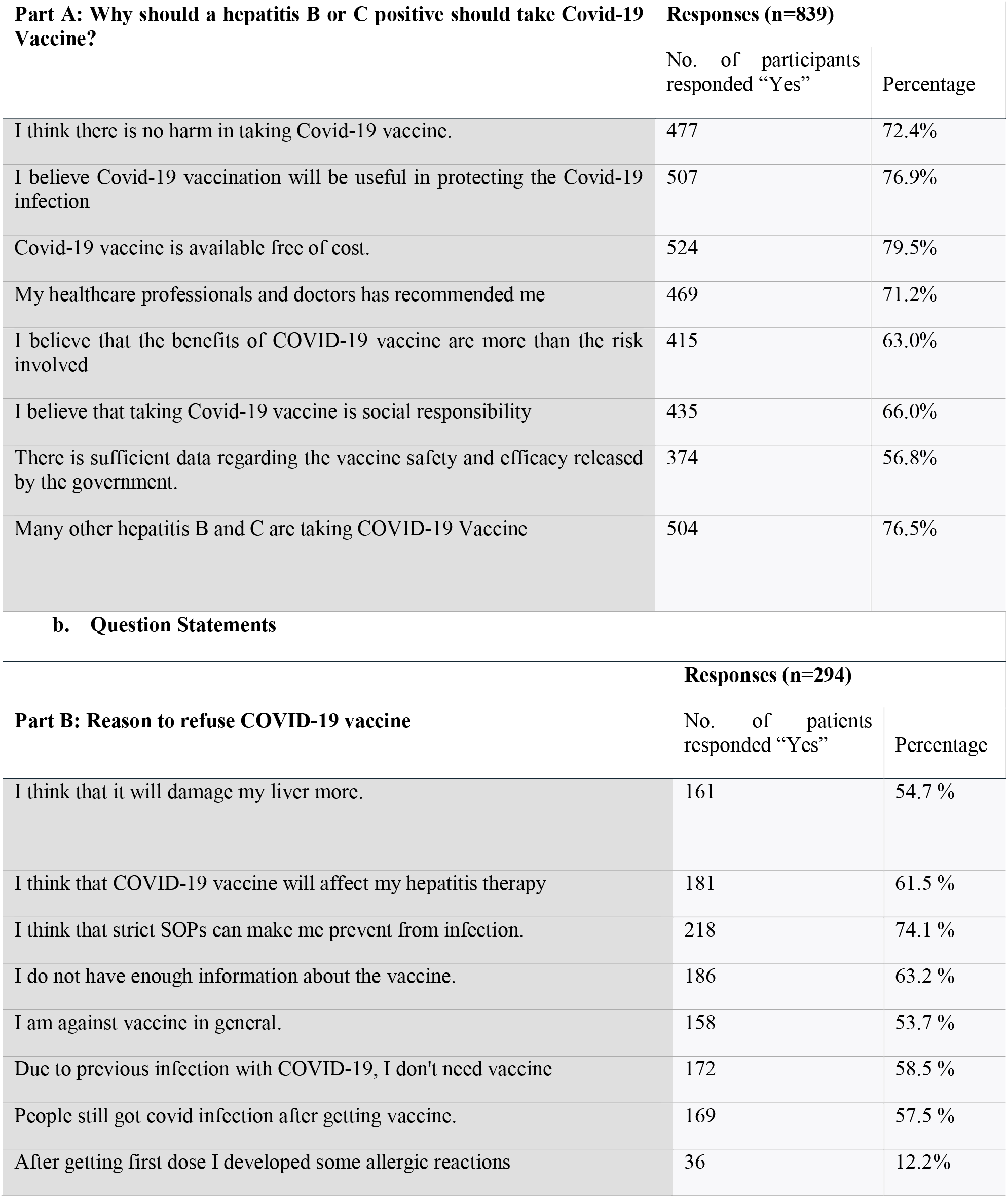
Assessment of patients choosing COVID-19 vaccine for protection. Table 2B. Question statements to assess Hepatitis patient for refusing COVID-19 vaccine.

| **Part A: Why should a hepatitis B or C positive should take Covid-19 Vaccine?** | **Responses (n=839)** |  |
| --- | --- | --- |
|  | No. of participants responded "Yes" | Percentage |
| I think there is no harm in taking Covid-19 vaccine. | 477 | 72.4% |
| I believe Covid-19 vaccination will be useful in protecting the Covid-19 infection | 507 | 76.9% |
| Covid-19 vaccine is available free of cost. | 524 | 79.5% |
| My healthcare professionals and doctors has recommended me | 469 | 71.2% |
| I believe that the benefits of COVID-19 vaccine are more than the risk involved | 415 | 63.0% |
| I believe that taking Covid-19 vaccine is social responsibility | 435 | 66.0% |
| There is sufficient data regarding the vaccine safety and efficacy released by the government. | 374 | 56.8% |
| Many other hepatitis B and C are taking COVID-19 Vaccine | 504 | 76.5% |

**b. Question Statements**
| **Part B: Reason to refuse COVID-19 vaccine** | **Responses (n=294)** |  |
| --- | --- | --- |
|  | No. of patients responded "Yes" | Percentage |
| I think that it will damage my liver more. | 161 | 54.7 % |
| I think that COVID-19 vaccine will affect my hepatitis therapy | 181 | 61.5 % |
| I think that strict SOPs can make me prevent from infection. | 218 | 74.1 % |
| I do not have enough information about the vaccine. | 186 | 63.2 % |
| I am against vaccine in general. | 158 | 53.7 % |
| Due to previous infection with COVID-19, I don't need vaccine | 172 | 58.5 % |
| People still got covid infection after getting vaccine. | 169 | 57.5 % |
| After getting first dose I developed some allergic reactions | 36 | 12.2% |

The questionnaire A analysis revealed that most participants (79.5%) were happy to get vaccinated because there were no charges of that vaccine as compared to the Hepatitis C vaccine available in the markets (Table 3a). Many participants (76.9%) believed that the vaccine would protect them from covid-19. About 76.5% people claimed that thy were encouraged when they realized that other hepatitis patients were also getting vaccine. As a reflection of their confidence in their doctors, 469 out of 710 people (71.2% of the total) vaccinated themselves after medical professionals encouraged them to do so. Yet 374 (56.8%) patients showed concern about the insufficient data available on the vaccine safety and efficacy published by the Public Health Department.

There were many explanations given by individuals when asked why they didn’t want to be vaccinated (Table 3b). About 74.1 % of participants had a view that precautionary measures like social distancing and wearing a mask or visor could protect them from getting coivd-19 infection. Some people refused (63.2 %) to get vaccination simply because they did not have enough information about the vaccine due to unavailability of literature regarding outcomes of covid vaccination. Interestingly, half of the patients (53.7 %) were simply against any type of immunization whatever vaccine it was and perceived the immunization as a part of foreign agenda. However, 181 (61.5 %) patients were worried that the vaccine might interact with their hepatitis therapy as there is no published data available which can prove it wrong; people don’t believe in assumptions especially in case of a health-related issue. The study also revealed that about 54.7 % of total participants were concerned about their liver condition as they were already dealing with a liver infection. The fear of an over-burdened liver cannot be overlooked. More than half (57.5 %|) of the patients who were against the vaccination were of view that there is no point to get vaccination if it is not protecting you against the infection. Around 58.5 % participants were of view that they have immunity against the disease due to the previous covid infection. Unfortunately, some patients (12.2 %) got allergic reactions after getting first dose due to which they feared to try another type of covid vaccine.

### Correlation between various parameters

Table 4a showed positive and negative correlation between various parameters of hepatitis patients with respect to the trend towards covid-19 vaccine. A very strong relationship (.104**) was found among the self-employed people toward the covid vaccine as compared to the people related to other sources of income. The trend showed that there was a significant difference among the people of various education levels. The participants having higher levels of education were more agreed to get vaccinated against covid-19 as compared to others. Surprisingly, participants having high incomes were showed a highly significant difference and were less likely to get vaccinated (−.119**) and same was the trend seen in case of females (−0.018). All age groups showed a weak relationship between patients and their attitude towards the covid vaccine. A negative correlation was seen in case of ethnicity and location. It was revealed that participants from Punjabi origin were reluctant (−0.064) to get vaccinated as compared to Kashmiri and Pathans. The study also revealed that less people from Muzaffarabad AJK were get vaccinated (−0.022) than those from Peshawar, KPK.

**Table 4a.** Correlation between different variables of Hepatitis B and C patients with the Covid-19 vaccine approval or rejection. b. Spearman correlation between different variables and COVID-19 vaccine.

| **a.** |  |  |  |  |  |  |  |  |  |  |  |
| --- | --- | --- | --- | --- | --- | --- | --- | --- | --- | --- | --- |
| Variables | Gender | education | location | income | Occupation | Ethnicity | Hepatitis B | Hepatitis C | Covid vaccine | COV 2nd dose | COV booster |
| Age | ↑ 0.124 | ↘ -0.126 | ↘ 0.03 | ↘ 0.016 | ↘ -0.03 | ↘ -0.004 | ↘ -0.126 | ↑ 0.126 | ↘ 0.033 | ↘ -0.015 | ↑ 0.07 |
| Gender |  | ↑ 0.064 | ↘ -0.055 | ↘ -0.095 | ↑ 0.057 | ↘ 0.025 | ↘ -0.08 | ↑ 0.087 | ↘ -0.018 | ↑ 0.093 | ↘ -0.046 |
| education |  |  | ↘ -0.137 | ↘ -0.041 | ↘ 0.023 | ↑ 0.05 | ↑ 0.081 | ↘ -0.081 | ↑ 0.09 | ↑ 0.182 | ↘ -0.005 |
| location |  |  |  | ↑ 0.058 | ↘ 0.009 | ↘ -0.034 | ↘ -0.06 | ↑ 0.06 | ↘ -0.022 | ↑ 0.048 | ↑ 0.094 |
| income |  |  |  |  | ↘ 0.044 | ↑ 0.095 | ↑ 0.075 | ↘ -0.075 | ↘ -0.119 | ↘ -0.106 | ↑ 0.083 |
| Occupation |  |  |  |  |  | ↑ 0.082 | ↘ 0.007 | ↘ -0.007 | ↑ 0.104 | ↑ 0.07 | ↑ 0.103 |
| Ethnicity |  |  |  |  |  |  | ↘ 0.03 | ↘ -0.03 | ↘ -0.064 | ↑ 0.083 | 0.044 |
| Hepatitis B |  |  |  |  |  |  |  | ↓ -1 | ↘ 0.041 | ↑ 0.096 | ↘ 0.006 |
| Hepatitis C |  |  |  |  |  |  |  |  | ↘ -0.041 | ↘ -0.096 | ↘ -0.006 |
| Covid vaccine |  |  |  |  |  |  |  |  |  | ↑ 0.394 | 0.028 |
| COV 2nd dose |  |  |  |  |  |  |  |  |  |  | ↑ 0.209 |
| **b.** |  |  |  |  |  |  |  |  |  |  |  |
| Variables | Spearman Correlation r |  |  |  |  |  |  |  | P Value (0.05) |  |  |
| Age | 0.033 |  |  |  |  |  |  |  | 0.340 |  |  |
| Gender | -0.018 |  |  |  |  |  |  |  | 0.612 |  |  |
| Education | 0.064 |  |  |  |  |  |  |  | 0.064 |  |  |
| Location | -0.022 |  |  |  |  |  |  |  | 0.522 |  |  |
| Income | -.119 |  |  |  |  |  |  |  | **0.001\*\*** |  |  |
| Occupation | .107 |  |  |  |  |  |  |  | **0.002\*\*** |  |  |
| Ethnicity | -0.035 |  |  |  |  |  |  |  | 0.309 |  |  |
| Hepatitis B | 0.041 |  |  |  |  |  |  |  | 0.240 |  |  |
| Hepatitis C | -0.041 |  |  |  |  |  |  |  | 0.240 |  |  |

### Relationship between Demographic Characteristics and Responses of Participants

Associations of different characteristics were made against the participants’ knowledge of covid-19 (Table 4b). First, education levels of the participants were evaluated against the use of mask by the participants. It was observed that the use of mask was significantly less among individuals with lower education levels as compared to individuals with higher education (P 0.064). However, non-significant relationships were present between education level and other aspects of our study such as attending social gatherings, frequent hand washing, and covid-19 as a bioweapon. Second, income levels of the participants were analyzed against various factors. A significant relationship was found among the income level of the participant and their affinity to join social gatherings during the covid-19 pandemic. It was observed that respondents with lower income levels were more likely to attend gatherings as compared to higher income respondents. Similarly, income level was also found to be associated with the subjects’ use of mask. Participants with higher income levels were wearing N95 and surgical masks, whereas cloth masks were usually used by lower-income respondents. Terminally higher- and lower-income category subjects wore fewer masks as compared to middle-income-range groups. More participants in the lower-income category had to leave their jobs or had their business shut down during the covid-19 pandemic as compared to higher-income respondents. Likewise, more women have left their jobs as compared to their male counterparts. However, business shutdown trend mainly affected the male gender. Moreover, the males were found to wear mask more often than the females. Females usually wore N95 masks, whereas male respondents relied more on surgical masks or cloth masks. On the contrary, no significant associations were present among all the characteristics with hand-washing practices and the use of covid-19 as bioweapons.

## DISCUSSION

In Pakistan, the combined number of persons infected with hepatitis B virus (HBV) and hepatitis C virus (HCV) is over 15 million^12^. Free treatment for viral hepatitis is being offered throughout the nation thanks to the Hepatitis Prevention and Treatment Program and other government-funded initiatives^23^. At a time when a worldwide effort to eliminate viral hepatitis was essential, covid-19 placed a significant burden on national healthcare systems. According to Mustafa et al.^24^ (2021), with the rise of covid-19 cases in early 2020, a sharp decline in visits of viral hepatitis patients was observed to their respective clinics across Pakistan. However, an estimated 325 deaths because of viral hepatitis were also reported each day. The data suggested that there was a clear shift of priority towards the Covid patients due to unavailability of a valid treatment and increased number of causalities.

Current study is unique as it primarily examined how the Hepatitis B and C patients residing in Peshawar KP and Muzaffarabad AJK have responded to the covid-19 immunization program. Currently no data is available regarding concerns of hepatitis patients in the area, therefore, we have assessed the participants concerns regarding the vaccine and compared it with the other studies conducted in the world.

As described in the Health Belief Mode, the preventive actions, like getting a vaccination, depend on how people see the risks of disease and how they perceive the effectiveness, benefits, and costs of these actions^25^. Pakistan and Afghanistan are two countries where nation-wide immunization has always been challenging as we have seen in the case of polio eradication program according to Shafique et al.^26^ (2021). In this study it was initially decided to contact maximum number of viral hepatitis patients residing in the study area. However, it was observed that most of the patients avoid such interviews due to some reasons. Therefore, we could succeed to interview around 839 patients. The survey revealed that majority of the participants (79.5%) were open to vaccination which shows appositive attitude towards the covid-vaccination. The reason might be the easy availability of the vaccine and an active covid-19 eradication program in which healthcare workers are going door to door to make sure everyone has got vaccinated. The results were even better than a similar study conducted by Tahir et al.^27^ in 2021.

However, there are still number of individuals which are not immunized due to some reason which is a real concern. It is also notable that majority of the individuals who participated in this study was between the age of 18 and (57.1 %) which indicates that young generation is more open to express their opinion about health issues. There were mixed responses when the participants were asked whether a patient suffering from viral hepatitis should get immunized.

Most of the patients (54.7 %) refused to get fully vaccinated despite the government’s strict orders because they were worried about their liver condition. Unfortunately, there is not much data about the effect of covid vaccines on the liver of hepatitis B and C patients specifically. Therefore, it is nearly impossible to persuade a person for getting a vaccination with a poor liver condition. Furthermore, there are some studies that have reported the postvaccination Autoimmune Hepatitis (AIH)^28-32^ which makes it essential to study the health of viral hepatitis patients after vaccination to devise a better strategy for hepatitis patients in order to prevent covid-19.

More than half of the patients (57.5 %) who refused to take the 2^nd^ dose individuals believed that covid-19 vaccination might not protect them from the covid-19 infection because they got ill or observed that people contracted infection even getting vaccinated. The concern is authentic; there are many cases reported around the world where vaccines have failed to prevent individuals from the disease. For instance, in a recent study Jain et al.^33^ (2021) reported that despite its effectiveness in preventing masses from the adverse effects of covid infection the covid vaccine was not proved suitable for certain individuals and postvaccination infection even deaths have been raised many questions on the efficacy of the vaccine especially in patients with vulnerability.

There were a few participants (12.2 %) who were covid against the vaccination because they developed adverse reactions after getting the 1^st^ dose of covid vaccine. This concern was not baseless as many though rare but incidents regarding hypersensitivity reaction to covid vaccine have been reported since 2020 vaccine roll out^34^.

About 218 (74.1 %) out of total participants believed that strict Standard Operating Procedures (SOPs) could prevent them from infection. Their belief was not wrong as we have seen that strict precautionary measures have saved masses when there was no vaccine available during the pandemic back in 2020. In random studies Kissler et al.^35^ (2020) and Moosa^36^ (2020) analyzed that social distancing has appeared to be quite effective to contain the covid infection when there was no vaccine available. Similarly, Hsieh et al^37^. 2020, reported that preventive measures such as social distancing, using Personal Protective Equipment (PPE), and frequent handwashing, etc. at a personal level could be significantly helpful to control the covid infection in the absence of other interventions. However, their approach to avoid covid-19 infection was not realistic. It is impossible to avoid social contact in this fast-paced life and sometimes SOPs are not enough. It is observed in various studies though minimal but there are still chances to get infected by following SOPs only^38^.

## CONCLUSION

It was concluded that despite a successful covid-19 prevention program in Pakistan the concern towards the safety and efficacy of covid vaccine is still there. The study revealed that the reason to refuse vaccination cannot be neglected as they are genuine and need to be addressed carefully. Additionally, addressing the issues regarding vaccine refusal requires genuine data demonstrating the benefits of vaccines in viral hepatitis patients is inevitable in order to resolve the ambiguity related to covid vaccination efficacy.

## Data Availability

will be provided on demand

## LIMITATIONS IN STUDY

Despite many efforts for making this study possible, it has a few limitations. First, the study area was confined to only two cities Peshawar and Muzaffarabad which is not enough to portray a clear picture of the current situation in Pakistan. Secondly, the resistance towards covid-19 vaccination-related reporting also makes data collection difficult. Natives especially hepatitis patients usually do not want to take part in such studies to avoid stigmatization.

## Footnotes

a https://www.nih.org.pk/wp-content/uploads/2020/04/COVID-19-NAP-V2-13-March-2020-1.pdf

b https://www.nih.org.pk/public/novel-coranavirus-2019-ncov

c https://covid19.who.int/region/emro/country/pk

d https://covid.gov.pk/

